# Could seasonal influenza vaccination influence COVID-19 risk?

**DOI:** 10.1101/2020.09.02.20186734

**Authors:** P. De Wals, M. Divangahi

## Abstract

**Background:** With possible resurgence of the SARS-CoV-2 and low seasonal influenza virus circulation next winter, reviewing evidence on a possible interaction between influenza vaccination and COVID-19 risk is important.

**Objective:** To review studies on the effect of influenza vaccines on non-influenza respiratory disease (NIRD).

**Methods:** Using different search strategies, 18 relevant studies were identified and their strength, limitations and significance were assessed.

**Results:** Analysis of 4 RCT datasets did not suggest increased NIRD risk in recipients of live-attenuated vaccines (LAIV) and results of a cohort study suggested short-term protection consistent with the hypothesis of ‘trained immunity’. One RCT, four cohort studies and one test-negative case-control suggested increased NIRD risk in recipients of inactivated influenza vaccines (IIV), whereas five test-negative case-control studies did not show an increased risk associated with a specific viral pathogen. Cross-protection against COVID-19 was suggested in one cross-sectional study on IIV but major biases could not be excluded. Results of four recent ecological studies on COVID-19 were challenging to interpret.

**Conclusions:** Available data on LAIV are reassuring but not all those on IIV. A drastic reorientation of 2020–2021 influenza campaigns is probably not warranted but studies aiming to test COVID-19 risk modification among recipients of seasonal influenza vaccines should be planned and funded.

## Introduction

In all high-income countries of the Northern hemisphere, immunization campaigns will be implemented this fall to reduce the burden of influenza disease, and eventually, to decrease ambulatory and hospital services use. In 2019, publicly-funded programs targeting high-risk groups were in place in British Columbia, New Brunswick and Quebec, whereas a universal vaccination approach was selected in the other provinces and territories in Canada (1). Similar strategies are expected to be renewed next fall. For the 2020–2021 season, the majority of influenza vaccine doses that will be distributed in Canada are tri-and four-valent inactivated influenza vaccines (IIV), including a high-dose formulation for use in adults > 65 years of age. A small quantity of the nasal live-attenuated influenza vaccine (LAIV) which is authorized for use in persons 2–59 years of age will also be distributed. Additionally, there is a IIV containing the MF59C.1 adjuvant that will eventually be available on the private market.

An increasing body of evidence suggests that live-attenuated vaccines such as the BCG vaccine, measles vaccine, smallpox vaccine and oral polio vaccine have non-specific beneficial protective effects against diseases caused by pathogens other than the targeted one (2). Conversely, inactivated vaccines such as the diphtheria-tetanus-pertussis vaccine or the inactivated polio vaccine have been associated with increased morbidity and mortality attributable to other causes (3). Both positive and negative ‘off-target’ effects of vaccines have been observed mainly in children and seem to be greatest for females than for males (4). These ‘off-target’ effects of vaccines are the result of epigenetical reprogramming of innate immune cells, called’trained immunity’ (2). Interestingly, different group of adjuvants used or tested in vaccines have also the capacity to enhance innate immune mechanisms that may induce unspecific broad-scale protection against infectious pathogens (5).

In the context of a persistent SARS-CoV-2 pandemic and a possible resurgence after the summer in the Northern hemisphere whereas the circulation of seasonal influenza viruses might be suppressed as observed since March in Australia (6), the safety of influenza vaccination is of special concern. The objective of this review was to assess evidence regarding a possible interaction between influenza vaccination and the risk of respiratory infections caused by other pathogens.

## Methodology

The literature search focused on human studies showing evidence of interference or interaction between influenza vaccination and the risk of disease caused by other respiratory pathogens, including SARS-CoV-2. The ‘snow ball’ technique was first used, starting with published manuscripts describing biological mechanisms of innate and trained immunity and reviewing evidence pertaining to vaccines. A first list of 13 relevant papers was identified. Thereafter, a systematic search in several databases using specific criteria was undertaken but no additional citation was found. Finally, the list of COVID-19 SARS-CoV-2 preprints from MedRxiv and BioRxiv was searched, bringing two additional citations. The list was supplemented by opportunistic Google searches and 3 recent and unpublished ecological studies were found, for a grand total of 18. Details on search strategies are shown in the supplementary material.

## Results

A total of 18 relevant manuscripts were identified, including 5 studies that were not peer-reviewed and accepted for publication in a scientific journal at the time of the writing of this manuscript (7–11). The focus, design, main methodological features and results of these 18 studies are shown in Table 1. Two studies focused on LAIV and 16 on unadjunvanted IIV, including 4 ecological studies on COVID-19. No study on high-dose IIV or adjuvanted IIV was found.

**Table 1:** Studies on the association between live-attenuated influenza vaccines (LAIV), inactivated influenza vaccines (IIV) and risk of respiratory disease other than influenza.

| Reference | Vaccine/Design | Methodological features | Main results |
| --- | --- | --- | --- |
| Piedra et al., 2007 (12). | LAIV<br>Community-based trial | An open-labeled, nonrandomized, community-based influenza vaccine trial was conducted in children 5 to 18 years of age in Texas. Eligible healthy children were offered a trivalent LAIV whereas children who had underlying risk conditions for influenza or who resided with immunocompromised household contact were offered a trivalent IIV. Vaccinations took place from October 10 to December 30, 2003. A total of 6 569 children received the LAIV and 1 040 the IIV. An influenza A (H3N2) outbreak occurred in the study community from October 12 to December 20, 2003. All residents in the community with febrile respiratory illness (FRI) were invited to have a throat culture. | During the outbreak, 55 episodes of FRI were identified in the LAIV group and 19 tested positive for influenza (34.5%), 24 episodes in the IIA group and 14 tested positive for influenza (58.3%), and 231 episodes in unvaccinated children with 127 positive tests (55.0%). Significant protection against influenza-positive illness was measured in the LAIV (37%; $p=0.006$) but no protection in the IIV group. Protection against influenza-positive illness in the LAIV group was observed during the first week following vaccine administration and lasted during the entire outbreak. protection against pneumonia and influenza clinical events was 50% In the LAIV group and non-existent in the IIV group. |
| De Serres et al. 2013 (13). | LAIV<br>Randomized control trials | Review of RCTs aiming to compare efficacy estimates of the live attenuated influenza vaccine (LAIV) in children $\leq 7$ years-old and the elderly $\geq 60$ years-old using either the classical *per-protocol* cohort analysis or the test-negative design (TND) in 4 datasets. During the influenza season, respiratory specimens were tested by viral culture and the primary outcome was culture-confirmed influenza (test-positive) due to any strain regardless of antigenic similarity. Specimens with influenza-negative culture were not further tested to identify other specific causative pathogens. | LAIV efficacy estimates and their confidence intervals were virtually identical for *per-protocol* RCT versus TND analyses, using three different control groups: (i) participants with an episode of non-influenza illness, no censoring for influenza; (ii) participants with an episode of non-influenza illness, with censoring for influenza; (iii) all non-influenza illness episodes. |
| Cowling et al., 2012 (17). | IIV<br>Randomized control trial | RCT in Hong Kong including 6-15-year-old children who received a trivalent inactivated influenza vaccine (n=69) or placebo (n=46) and were observed during 9 months in 2008-2009. | TIV recipients had an increased risk of virologically-confirmed non-influenza infections: relative risk: 4.40 (95%CI: 1.31 to 14.8). |
| Andersen et al., 2018 (18). | IIV<br>Cohort study | In October 2010, a national campaign with an inactivated influenza vaccine was implemented in Guinea-Bissau, targeting children 6-60 months of age. The target population included 3 cohorts of children who participated in different clinical trials. Age-adjusted all-cause mortality rate ratios (MMR) in experimental and control groups before and after the influenza campaign were computed. | The H1N1 campaign was associated with an increase in mortality: MRR = 1.86 (1.02 to 3.42). This effect was only found for girls (MMR = 2.32; 95%CI: 1.19 to 4.52), not for boys (MMR = 0.95: 0.23 to 3.92). |
| Hansen et al., 2019 (19). | IIV<br>Cohort study | Medical consultations in a cohort of 9 862 children 6-59 months of age in Guinea-Bissau were recorded. In October 2010, a mass immunization campaign with inactivated unadjuvanted influenza pandemic H1N1 vaccine was implemented. Consultations rates before and after the H1N1 immunization campaign were compared by proportional hazard ratios | Among 10 290 children eligible for the H1N1 campaign, 60% had been vaccinated, 18% had not and for 22% no information was available. After the H1N1 campaign, the consultation rates tended to decline less for vaccinated children (HR = 0.80; 95%CI: 0.75 to 0.85) than for non- participants (HR = 0.68; 95%CI: 0.58 to 0.79; $p = 0.06$ for same effect between groups). |
|  |  | (HRs) using a Cox mode, adjusting for seasonality, maternal education, ethnicity and residential district in stratified analyses. |  |
| Dierig et al., 2014 (20). | IIV Cohort study | In Australia, 399 children 6-59 months of age were prospectively recruited from child care centers and classified as fully vaccinated, partially vaccinated and unvaccinated according to their receipt of unadjuvanted A (H1N1)pdm09 vaccine in 2010. Parents were regularly contacted for the occurrence of influenza-like illness (ILI) during the March-August period. Nose and/or throat swabs were taken for multiplex respiratory virus PCR testing. | Influenza-vaccinated children (n=143) were 1.6 times (P = 0.001) more likely than unvaccinated children (n= 238) to have a non-influenza ILI (37.1% vs 24.8%; p=0.001) or ILI caused by virus other than influenza (32.3% vs 21.8%; p=0.02). |
| Rikin et al., 2018 (21). | IIV Cohort study | A prospective study among 250 household in New York City was conducted during the period 2013-2016, encompassing 3 influenza seasons (MoSAIC community surveillance study). Influenza vaccination was confirmed through city or hospital registries. Cases of acute respiratory illness were ascertained by twice-weekly text messages to household. Nasal swabs were obtained from ill participants and analyzed for respiratory pathogens using multiplex PCR. The primary outcome measure was the hazard ratio (HR) of laboratory-confirmed ARI in the post-vaccination period compared to other time periods during three influenza seasons. | The analysis included 687 children and 312 adults. The hazard of non-influenza respiratory pathogens was higher during the 14-day post-vaccination period (HR 1.65, 95% CI [1.14, 2.38]); when stratified by age the hazard remained higher for children (HR 1.71; 95% CI: 1.16, 2.53) but not for adults (HR 0.88; 95% CI: 0.21 to 3.69). |
| Kelly et al., 2011 (22). | IIV Test-negative design | A prospective study in general practices and a hospital emergency department among children 6-59 months of age in Sydney, Australia, in 2008-2009. Tri-valent inactivated influenza vaccine effectiveness (VE) against laboratory-confirmed influenza was estimated with cases defined as children with an influenza-like illness who tested positive and controls as those with an influenza-like illness who tested negative for influenza virus (total = 289 children). | Using all the influenza-negative controls, the adjusted VE was 58% (95%CI: 9 to 81). When controls were limited to those with another virus present, the adjusted VE was 68% (95%: 26 to 86). |
| Sundaram et al., 2013 (23). | IIV Test-negative design | The association between influenza vaccination, and detection of respiratory viruses among 1616 children <5 years old and 1568 adults ≥50 years old with acute respiratory illness was investigated, using the test-negative method. Nasopharyngeal samples collected from 2004–2005 through 2009–2010 in the US were tested for 19 respiratory virus targets using RT-PCR. Vaccination status was determined using a validated registry. | Non-influenza respiratory viruses were detected in 70% of 2010 children and 38% of 1738 adults without influenza. The proportion vaccinated did not vary between non-influenza virus-positive controls and pan-negative controls in children (p = 0.62) or adults (p = 0.33). |
| Blyth et al., 2014 (24). | IIV Test-negative design | An observational study enrolling children with influenza-like illness presenting to a tertiary-care pediatric hospital in Australia was conducted in 2008–2012. Vaccination status was determined by parental questionnaire and confirmed via the national immunization register and/or vaccine providers. Respiratory virus PCR and culture were performed on nasopharyngeal samples. The test-negative design was used to estimate vaccine effectiveness (VE) by using 2 control groups: | Of 2 001 children enrolled, influenza was identified in 389 and another respiratory virus in 1134 children. Using test-negative controls, VE was 64.7% (95%CI: 33.7 to 81.2) and 65.8 with OVD controls (95%CI: 32.1 to 82.8). |
|  |  | all influenza test-negative subjects and other-virus-detected (OVD) subjects. |  |
| Feng et al., 2017 (25). | IIV<br>Test-negative design | From 2010 to 2013, outpatients with acute respiratory illness presenting to clinics in 13 US jurisdictions with acute respiratory infections were tested for influenza and other respiratory viruses. Self-reported vaccination status, demographic data, and clinical data were collected by general practitioners at each study site. Influenza vaccine effectiveness was estimated using three different control groups: patients testing negative for influenza virus, patients testing negative for influenza virus but positive for at least one other respiratory virus, and patients testing pan-negative (those who tested negative for both influenza and other respiratory viruses). | Based on 10 650 patients 6 months to 99 years of age, no difference in estimates of vaccine effectiveness against any influenza virus when using influenza test negative controls, other respiratory virus positive controls, or pan-negative controls. No association was found between influenza vaccination and detection by age group of any other respiratory virus including RSV, rhinoviruses, PIV 1-3, MPV and adenovirus. |
| Skowronski et al., 2020 (26). | IIV<br>Test-negative design | Inactivated influenza vaccine effectiveness against influenza and non-influenza respiratory viruses (NIRV) was assessed by test-negative design using historic datasets of the community-based Canadian Sentinel Practitioner Surveillance Network (SPSN), spanning 2010-11 to 2016-17. | The study included 4281 influenza, 2565 NIRV, and 3841 pan-negative specimens. The OR for influenza vaccination among influenza cases versus influenza test-negative controls was 0.55 (95%CI=0.50 to 0.61), corresponding to a VE of 45% (95%CI=39 to 50%). ORs were similar with pan-negative (0.58; 95%CI=0.52 to 0.65) or NIRV-positive controls (0.51; 95%CI=0.45 to 0.58). |
| Wolff, 2020 (27). | IIV<br>Test-negative design | The vaccination status of US military personnel with laboratory-confirmed influenza (N= 3349), non-influenza respiratory viruses (N=2880) and negative result (N=3240) during the 2017-2018 influenza season were compared. | Comparing vaccinated to non-vaccinated patients, the adjusted odds ratio for non-influenza viruses was 0.97 (95% CI: 0.86 to 1.09; p = 0.60). Examining non-influenza viruses (NIV) specifically, the odds of both coronavirus and human metapneumovirus in vaccinated individuals were significantly higher when compared to unvaccinated individuals (OR = 1.36 and 1.51, respectively). |
| Finck et al., 2020 (8). | IIV<br>Cross-sectional study | A total of 92,664 clinically- and molecularly-confirmed Covid-19 cases reported in the Brazilian surveillance system for severe respiratory infections between January 29, 2020 and June 8, 2020. All health facilities and providers throughout the country are invited to report all infections disease cases into a central system using a standardized reporting protocol. The primary outcome was patient survival and secondary outcomes were intensive care treatment and invasive respiratory support. The influenza immunization status of the patient is one of the variables on the reporting form. In Brazil, trivalent IIV are exclusively used and offered free of charge to individuals 60 years and older, to individuals less than 60 years with high-risk condition and to health care workers. The 2020 annual influenza vaccination campaign was launched in late March, one month earlier than originally planned, to ensure vaccine delivery to the public prior to the incoming wave of SARS-CoV-2 infections, and with the ambition to reach a total of 67.6 M people in a population of 2010 M. | Patients who received a recent influenza vaccine experienced on average 8% lower odds of needing intensive care treatment (95%CI: 0.86 to 0.99), 18% lower odds of requiring invasive respiratory support (95%CI: 0.74 to 0.88) and 17% lower odds of death (95%CI: 0.75 to 0.89). Lower risks were observed in all age-groups and also in sensitivity analyses restricting, stratifying or adjusting for age, gender, educational attainment, ethnicity, pre-existing risk conditions and the health care facility. |
| Arokiaraj et al., 2020 (7). | Ecological study | Correlations between influenza vaccine coverage statistics in persons $\geq 65$ years and COVID-19 morbidity/mortality data extracted from the Coronavirus-Worldometer were performed, adjusting or not for the intensity of testing in 35 high- and middle-income countries. | Weak negative correlation coefficients were found but no statistical test was performed. |
| Lisewski, 2020 (9). | Ecological study | COVID-19 infections rates in 29 OECD countries were correlated with influenza vaccine coverage in the 65 years and over age-group using published statistics | In the main analysis, Pearson correlation coefficient $r$ between influenza vaccine rates and the attack rates for these 29 OECD countries was $r = 0.58$ (95CI: 0.27 to 0.78; $p=0.001$), suggesting an increased risk associated with vaccination. |
| Zanettini et al., 2020 (10). | Ecological study | COVID-19 mortality data between January 22, 2020 and June 10, 2020 in 2 034 counties across the 50 US states and Washington D.C. were analyzed according to influenza vaccination coverage data in the population aged 65 and older. A series of county-specific demographic, socio-economic, health, health care and environmental variables were included in multivariate statistical models and a propensity score was constructed. | Results suggested a (non-linear) reduction in COVID-19 mortality associated with higher influenza vaccination rates in the elderly population, with a statistically-significant 28% decrease in the COVID-19 death rate for a 10% increase in vaccination coverage. |
| EBMPHET Consortium, 2020 (11). | Ecological study | Influenza vaccine coverage data in persons $\geq 65$ years of age in 35 countries of Europe and 49 states in the US were correlated with data on COVID-19 deaths and cases as of 22 May 2020. | Statistically-significant positive correlations between vaccine coverage rates (VCR) and reported COVID-19 incidence and mortality were found in Europe and the US. A statistically-significant positive correlation was also found between the VCR and the COVID-19 case fatality rate for Europe and the correlation was not statistically significant for the US. |

### Studies on live-attenuated influenza vaccines

The first study focusing on LAIV was a community-based trial in Texas that was launched at the same time of an influenza outbreak in late 2003. The outbreak was caused by an A (H3N2) strain that was poorly matched to the reference strain included in 2003–2004 seasonal LAIV and IIV vaccines (12). All children 5–18 years of age in the community were invited to receive a trivalent LAIV and those with a risk condition or who resided with immunocompromised household contact were offered a trivalent IIV. The simultaneous start of the campaign and the outbreak allowed estimation of vaccine effectiveness from the date of administration of the first dose. Results showed 37% protection in LAIV recipients starting from the first week after vaccine administration, which was statistically-significant. No protection was observed in IIV recipients. Protection was also observed for specific clinical outcomes, suggesting that reduced viral shedding in the LAIV group without any clinical benefit is not a likely explanation. Although no serology test was performed, the authors concluded that early protection was attributed to innate immune mechanisms triggered by the LAIV.

A study by De Serres and coworkers was carried out to compare efficacy estimates of LAIV derived from the classical per-protocol analysis of randomized clinical trials and those obtained by a test-negative design (TND) analysis using four datasets (13). TND is a modified and efficient version of the classic case-control study design in which test-positive results are cases and test-negative results are controls (14). Results showed that the LAIV efficacy estimates and their confidence intervals were virtually identical for per-protocol RCT versus TND analyses using three different computation methods. The authors concluded that a core assumption of the TND approach being that the intervention (LAIV in this case) has no effect on other non-targeted etiologies resulting in similar clinical outcome was confirmed. We reanalyzed the data included in Figure 2 of De Serres’ manuscript applying the per-protocol RCT cohort analysis to compute VE estimates against presumed non-influenza acute respiratory illnesses, we categorized patients with a least one influenza-negative result or all influenza-negative specimens (Table 2). Relative risk/rate estimates were slightly inferior to unity in 7 out of 8 calculations. The sensitivity of viral cultures to identify influenza in throat or nasopharyngeal swabs is not perfect but there is no gold standard to assess this precisely. RT-PCR seems to be a more sensitive diagnostic test that viral culture and comparisons were made in two studies (15,16). While in patients less than 40-year-old, detection rates by RT-PCR were 15–25% higher than detection by culture, the detection rates were very similar in older adults. In order to evaluate the impact of this factor on the risk of non-influenza illnesses associated with LAIV, sensitivity analyses were performed (Table 2). Assuming a 75% sensitivity of viral culture from clinical specimens, relative rates remained close to unity. To double the risk of disease caused by other pathogens (i.e. RR = 2), the sensitivity of viral culture for influenza would have to be around 17%. All together, these results suggest that LAIV given during the fall does not increase the risk of all non-influenza acute respiratory illnesses during the winter season.

**Table 2:** Frequency of persons with at least one negative culture for influenza and rate of influenza-negative specimens in four datasets from randomized clinical trials on the life-attenuated influenza vaccine (LAIV) (Source: DeSerres et al., 2013).

| Reference | Study population | Outcome | LAIV Group |  |  | Control Group |  |  | Relative Rate | 95% confidence Interval | Adjusted relative rate * |
| --- | --- | --- | --- | --- | --- | --- | --- | --- | --- | --- | --- |
|  |  |  | No participants | No outcomes | Rate | No participants | No outcomes | Rate |  |  |  |
| Belshe et al., 1998 (42). | Children 15-71 months of age<br>Season 1 | At least one influenza-negative culture | 1070 | 851 | 0.795 | 532 | 424 | 0.797 | 0.998 | 0.947 to 1.052 |  |
|  |  | Influenza-negative specimens | 1070 | 1959 | 1.831 | 532 | 939 | 1.765 | 1.037 | 0.959 to 1.122 | 1.072 |
| Belshe et al., 2000 (43).. | Children 26-85 months of age<br>Season 2 | At least one influenza-negative culture | 917 | 596 | 0.650 | 441 | 298 | 0.676 | 0.962 | 0.888 to 1.042 |  |
|  |  | Influenza-negative specimens | 917 | 1181 | 1.288 | 441 | 575 | 1.304 | 0.988 | 0.893 to 1.093 | 1.016 |
| Lunn et al., 2010 (44).. | Children 11-23 months of age | At least one influenza-negative culture | 819 | 708 | 0.864 | 413 | 372 | 0.901 | 0.960 | 0.920 to 1.001 |  |
|  |  | Influenza-negative specimens | 819 | 2889 | 3.527 | 413 | 1566 | 3.792 | 0.930 | 0.875 to 0.990 | 0.935 |
| De Villers et al., 2009 (45). | Adults ≥60 years of age | At least one influenza-negative culture | 1620 | 944 | 0.583 | 1622 | 981 | 0.605 | 0.964 | 0.910 to 1.020 |  |
|  |  | Influenza-negative specimens | 1620 | 1847 | 1,140 | 1622 | 1880 | 1.159 | 0.984 | 0.922 to 1.050 | 0,994 |

\*Adjusted assuming 75% sensitivity of viral culture from throat/nasopharyngeal specimens to identify influenza virus.

### Studies on inactivated influenza vaccines

#### Study 1

In a randomized clinical trial (RCT) on a trivalent IIV for children in Hong-Kong, vaccine recipients had shown a significant increase in risk of virologically-confirmed non-influenza infections during the 2008–2009 season compared to the placebo group (relative risk = 4.40; 95%CI: 1.31 to 14.8). Non-influenza virus infection was a secondary outcome in this RCT involving a small number of participants: 69 in the IIV group and 46 in the placebo group (17). A selection or information bias can be reasonably excluded in this study but not a random variation between groups in a RCT with a small sample size.

#### Study 2–3

Two cohort studies were conducted among children in Guinea-Bissau who were exposed to a mass immunization campaign using an unadjuvanted H1N1 (2009) pandemic IIV in October 2010 (18,19). Although influenza surveillance was not conducted in Guinea-Bissau, data from the neighboring country, Senegal, indicated that H1N1 influenza was circulating in the region at that time. In the first investigation (18), age-adjusted all-cause mortality rate ratios (MRRs) were evaluated before and after the influenza campaign in three cohorts of children who were enrolled with similar inclusion criteria in three clinical trials of BCG, OPV and vitamin A supplementation. Results showed an increased MMR after the influenza vaccination in the three cohorts with a combined effect of 1.86 (95%CI: 1.02 to 3.42), suggesting an increased mortality risk associated with IIV exposure. Importantly, this effect was present in girls only. The second investigation focused on all-cause medical consultations among 6 to 59-monthold children who resided in a district of the Guinea-Bissau capital city selected for active surveillance of different health outcomes (19). Consultation rates in vaccinated and unvaccinated children over time were compared by a Cox proportional hazard (HR) model. After the influenza vaccination, consultation rates tended to increase among vaccinated children (HR = 0.80; 95%CI: 0.75 to 0.85) than among unvaccinated children (HR = 0.68; 95%CI: 0.58 to 0.79), suggesting a negative ‘off-target’ effect of IIV. In these two investigations, specific causes of mortality or medical consultation were not investigated but respiratory infections certainly played a major role.

#### Study 4

In a cohort study conducted in Australia, 6–59-month-old children were prospectively recruited from childcare centers and classified according to receipt of the inactivated unadjuvanted A (H1N1) pandemic vaccine in 2010. Parents were regularly contacted for the occurrence of influenza-like illness (ILI) and nose/throat swabs were obtained from sick children for multiplex PCR testing (20). Vaccinated children were 1.6 times (p = 0 001) more likely than unvaccinated children to have a non-influenza illness(37.1% vs 24.8%; p = 0.001) or ILI caused by virus other than influenza (32.3% vs 21.8%; p = 0.02). A bias caused by a differential health care seeking behavior of parents can be reasonably excluded in this study but not a ‘healthy vaccinee’ effect.

#### Study 5

A prospective cohort study was conducted in a sample of households in New York City during the period 2013–2016 (21). Influenza vaccination status of participants was confirmed through city or hospital registries. Cases of acute respiratory illness were ascertained by twice-weekly text messages. Nasal swabs were obtained from ill participants and analyzed for respiratory pathogens using multiplex PCR. A 14-day post-vaccination risk period was chosen to assess the association of vaccination with the induction of influenza-specific immunity and its contributions to other potential pulmonary infections. The risk of non-influenza respiratory pathogens during this 14-day window was higher among vaccinated individuals compared to non-vaccinated individuals (Hazard Ratio = 1.65; 95%CI: 1.14 to 2.38); when stratified by age the hazard remained higher for children (HR = 1 71; 95% CI: 1.16 to 2.53) but not for adults (HR =0.88; 95% CI: 0.21 to 3.69). Many biases can be excluded in this study as a result of the self-controlled nature of the comparison.

#### Studies 6–11

There were 6 studies based on respiratory virus surveillance data analyzed by the test-negative design (TND) which is a modified and efficient version of the classic case-control study design. In TND, test-positive results are cases and test-negative results are controls and an Odds Ratio (OR) is computed to approximate a Relative Risk (RR) leading to a vaccine effectiveness estimate (14).

In a first TND study among 289 children seen in general practices and a hospital emergency department in Sydney, Australia, in 2008–2009, IIV effectiveness was 58% using all the influenza-negative controls, whereas the estimate was 68% when controls were limited to those with another virus present, suggesting interaction between influenza vaccine status and non-influenza disease risk (22).

No apparent interaction was reported in four other TND studies on IIV based on large sample sizes: (i) one study among children and adults in a cohort of patients seen by family physicians in the US (23), (iii) a study among children seen in a tertiary-care hospital in Australia (24), (iii) a study among outpatients seen in 13 surveillance sites in the US (25), and (iv) a study among patients seen by family physicians participating in the Canadian Sentinel Practice Surveillance Network (26). In two of these studies, no significant imbalance in OR was observed for respiratory syncytial virus, rhinovirus, human metapneumovirus, parainfluenza viruses and seasonal coronaviruses, suggesting no specific increased or decreased risk for these viruses associated with IIV administration (25,26).

Another highly-publicized manuscript being used by organizations skeptical of vaccines was authored by Wolff in January 2020 (27). This study described the distribution of respiratory virus results among Department of Defense personnel according to their past influenza vaccination status. In one of the sub-group analyses, the odds of both coronavirus and human metapneumovirus in vaccinated individuals were significantly higher when compared to unvaccinated individuals. The study was criticized on several grounds (26,28). First, influenza test results were reported from different sources and there was no standardization in testing. Second, results were not adjusted for seasonality and it is well known that different viruses are circulating at different times in the year. Third, in the unadjusted analysis of specific non-influenza viruses, including coronaviruses, influenza test-positive specimens were included in the test-negative control group for non-influenza viruses. In the context of effective influenza vaccine, influenza cases would have a lower likelihood of vaccination and as such, their inclusion would systematically reduce the proportion vaccinated in the control group, artificially inflating ORs comparing vaccine exposure between non-influenza cases and controls. In a re-analysis of Wolff’s data, Skowronski and coworkers showed that by excluding influenza-positive specimens in the control groups, the OR for coronavirus-positive illness for IIV recipients was 1.17 (95%CI: 0.97 to 1.40), instead of 1.44 (95%CI: 1.20 to 1.72) in the original analysis (26).

In TND studies, clinical specimens from oral/nasopharyngeal swabs are tested by multiplex RT-PCR and/or cultures and results classified into 3 categories: positive for influenza, negative for influenza and positive for another virus, negative for all viruses tested. A basic assumption of the TND is that the risk of disease caused by the non-influenza viruses is not modified by the influenza vaccine status (14). When this condition is met, VE estimates derived from the use of influenza-negative tests, influenza-negative but other virus-positive tests or pan-virus-negative tests as a control group are similar and unbiased compared with results obtained from a classic cohort design using an odds ratio for the measure of disease risk. Of note, viral cultures and multiplex RT-PCR kits are not 100% sensitive in the field and not all respiratory pathogens are tested, meaning that a uniform increase in respiratory disease risk across all respiratory pathogens would not be detected in a study based on the test-negative design. This is exactly what would be expected from ‘trained immunity’ mechanisms leading to hypo-responsiveness (2)

#### Study 12

Recently, a cross-sectional study (not peer-reviewed at the time of writing of this manuscript) of Brazilian surveillance data for severe respiratory infections was conducted to explore the association between COVID-19 severity and recent influenza vaccination (8). Results showed an 8% lower risk of intensive care admission, a 18% lower risk of invasive respiratory support care and a 17% lower risk of death in IIV recipients. According to Worldometer (as of July 15, 2020), the first COVID-19 case was identified on February 25, 2020. Disease incidence increased sharply in early May and by June 8, 2020, about 711 000 cases were reported (compared with 93 000 cases in Finck’s study) with no sign of slowing down. The epidemic put tremendous pressure on the health system including its administrative component. In this study, the completeness of COVID-19 diagnosis and reporting and the quality of data on immunization status and potential confounders were not assessed. Importantly, this study indicated that patients who received the influenza vaccine after COVID-19 symptom onset were equally if not better protected than those who were vaccinated before disease onset demands certainly further investigations.

### Ecological studies on COVID-19

Finally, four recent ecological studies (not peer-reviewed at the time of writing of this manuscript) on the association between IIV vaccine uptake in people ≥ 65 years of age and COVID-19 were identified. The first study focused on 35 high-and middle-income countries worldwide and concluded that influenza vaccination may provide protection against COVID19 (7). The second study focused on 29 OECD countries and concluded that influenza vaccination may be a risk factor (9). In these two studies, crude methodologies were applied and important confounding factors related to the quality of COVID-19 data and the timing of the epidemic at each country were not considered. However, a more robust analysis was conducted in the US influenza vaccination coverage in the population > 65 years of age, adjusting for a series of county-specific demographic, socio-economic, health, health care and environmental variables (10). These results suggested a significant reduced COVID-19 mortality among influenza vaccinated people (28% decrease in the COVID-19 death rate for a 10% increase in vaccination coverage). The study was restricted to COVID-19 deaths reported up to June 10, 2020, before the spread of the epidemic in Southern states and more rural areas. In the last study, statistically-significant positive correlations were found between vaccine coverage rates and reported COVID-19 incidence and mortality were found in Europe and the US (11). COVID-19 data up to May 22, 2020, were analyzed and no confounding variable was considered. In all these ecological analyses, inferences about individual risk that are based on inferences about the group to which they belong should be made with great care as unmeasured confounders may be present, leading to a false inverse relationship, the ‘ecological fallacy’ (29).

## Discussion

In this review, the re-analysis of 4 RCT datasets did not suggest any increased risk of non-influenza respiratory infection in LAIV recipients and the rapid induction of nonspecific protection against viral infection was observed in one prospective community trial (Table 1). Conversely, an increased risk of non-influenza respiratory infection among IIV recipients was suggested in one RCT, four cohort studies and one case-control study using a test-negative comparison (Table 1). In five other observational studies applying a test-negative design, influenza vaccine effectiveness estimates were not influenced by the choice of the control group, suggesting no specific interference with a particular pathogen although the hypothesis of a uniform risk modification across different non-influenza pathogens cannot be excluded. A protective effect of IIV against COVID-19 infection was suggested in a cross-sectional study presenting major sources of bias. Researchers and editors of scientific journals are always interested in intriguing observations and a publication bias cannot be excluded. Nevertheless, all the studies included in this review merit consideration.

Influenza infection triggers both innate and adaptive immune responses (30). In animal models, influenza virus infection was shown to provide rapid and extended protection against other viral infections (31,32). Several innate defense mechanisms are involved in unspecific host defense following influenza infection, including Type I interferons, tumor necrosis factor-α, neutrophils, macrophages, monocytes, dentritic cells and natural killer cells (31–35). LAIV administered intra-nasally mimics a wild influenza virus infection in which protective innate immune mechanisms are triggered (36). All these observations coupled with results of the present review are reassuring as to the safety of LAIV in a context of SARS-CoV-2 circulation.

Split and subunit IIV currently distributed have been designed to preferentially target adaptive immunity and enhance the production of antigen-specific protective antibodies(37). In animal and human studies, the protective effects of IIV was less than LAIV against heterologous influenza strains (35,36,38). While LAIV has the capacity to target innate immunity by inducing the expression of several interferon-related genes, IIV induced mainly a signature of genes that highly expressed in adaptive immunity and particularly in plasma B cells (39). Although *in vitro* studies have shown that seasonal IIV has the ability to reprogram myeloid and natural killer cells with enhanced capacity to produce anti-viral cytokines (40), the *in vivo* translational aspect of this study remains to be determined. How IIF could increase the risk or severity of a heterologous viral infection is not clear. Enhanced trained immunity often leads to reduce viral load and thus decrease magnitude of inflammatory responses and subsequently tissue damage. Thus, the initial control of viral replication will have a profound impact on the severity of disease which is largely dictated by inflammatory responses. If the viral multiplication is not reduced, enhanced trained immunity may reveal or exacerbate clinical symptoms, the ultimate expression being the cytokine storm (41). An adverse effect of IIV vaccination in a context of SARS-CoV-2 infection is currently a theoretical possibility and thus certainly required further investigation.

## Conclusion

Available data on LAIV are reassuring but not all those on IIV. Considering the limited amount and quality of evidence for an adverse effect of influenza vaccination on other respiratory viral infections, it would be difficult to suggest a suspension or a drastic change in influenza vaccination campaigns which are planned for the fall of 2020. A practical proposal would be to recommend LAIV for the immunization of high-risk children where feasible. Furthermore, we urgently need to plan and fund both fundamental and clinical studies aiming to investigate the potential impact of influenza vaccination on COVID-19 risk and severity.

## Data Availability

all data referred to in the manuscript are in the public domain and available as stated in the references

## Acknowledgements

The authors thank Maria-Eugenia Espinoza-Moya, PhD student at the University of Toronto, for assistance in the literature search and Benoît Soubeyrand, consultant at Blossom Vaccinology, for useful comments on the manuscripts.

## Contribution

PDW and MD contributed equally to the review and writing of the manuscript

